# Diversity and level of evidence evaluation of commercial pharmacogenomic testing for mental health

**DOI:** 10.1101/2022.11.07.22282051

**Authors:** José J. Morosoli, Penelope A. Lind, Kristina Spears, Gregory Pratt, Sarah E. Medland

## Abstract

This study examined arrays offered by commercial pharmacogenomic (PGx) testing services for mental health care in Australia and the United States, with a focus on utility for non-European populations. Seven of the 14 testing services we identified provided the manifests of their arrays. We examined allele frequencies for each variant using data from the Allele Frequency Aggregator^1^ (ALFA), genome Aggregation Database^2^ (gnomAD), Exome Aggregation Consortium^2^ (ExAC), and Japanese Multi Omics Reference Panel^3^, and examined genetic heterogeneity. We also analyzed meta-data from the Pharmacogenomic Knowledge Base^4^ (PharmGKB) and explored the biogeographical origin of supporting evidence for clinical annotations. Most arrays included the minimum allele set recommended by Bousman et al^5^. However, few arrays included *HLA-A* or *HLA-B*. The most diverse allele frequencies were seen for variants in *CYP3A5, ADRA2A* and *GNB3*, with European and African populations showing the largest differences. Most evidence listed in PharmGKB originated from European or unknown ancestry samples.

## Introduction

Genetic heterogeneity poses one of the main challenges to the translation of PGx findings in non-European populations and there is a high risk that it will lead to growing health disparity between European and non-European communities in precision medicine^6^. Since 1960, more than 10,000 publications with the terms “pharmacogenomic” or “pharmacogenetic” in their tile or abstract have been indexed by PubMed, with an average of 661 new publications per year since 2010, but only 222 publications included the terms “ancestry”, “heterogeneity”, or “ethnicity”. Despite the rising availability of commercial PGx services, previous research using ExAC data found that half of all the functional variants in drug-related genes are unique to only one of six populations (i.e., African/African American, South Asian, East Asian, Finnish and Non-Finnish European), including genes involved in the metabolization of psychoactive medication. Furthermore, only 0.1% of functional variants occur with an allele frequency ≥1% across ancestries^7^. Data from the 1000 Genomes Project showed significant inter-population minor allele frequency differences in 159 drug-response related SNPs across populations^8^. An exploration of PGx allele and phenotype frequencies for 487,409 participants in the UK Biobank showed that non-European populations carry a greater frequency of PGx variants that are predicted to be functionally deleterious^9^. Utility of the genetic variants included in mental health PGx testing services across ancestries is, therefore, unclear, especially considering that approximately one-fifth of new medications approved by the US Food and Drug Administration from 2008 to 2013, showed differences in exposure and/or response across racial/ethnic groups, translating to population-specific prescribing recommendations^10^.

Despite this uncertainty, in recent years there has been an increase in the number of peer reviewed articles supporting prophylactic PGx testing and its use in general practice^11^. A number of recommendations have been made about the quality and adequacy of commercial PGx genotyping arrays for mental health^5,12,13^. For example, PGx testing for two cytochrome P450 genes (i.e., *CYP2D6, CYP2C19*) is recommended to inform selection and dosing of antidepressant and antipsychotic medication^14^. However, key barriers to the implementation of PGx testing in psychiatry has been the lack of consensus on which *CYP2D6* and *CYP2C19* alleles were being tested, lack of large-scale replication studies, and flaws in study designs^13-15^. In view of the increasing availability of PGx testing for mental health, both to health professionals and directly to consumers, the need to compare the coverage of the arrays used by PGx services, and the potential differential utility of arrays across ethnic groups is paramount.

In the present study we aimed to examine: (i) the current composition of arrays offered for PGx testing for psychiatry in Australia and the US; (ii) differences in prevalence of these key PGx genetic variants between European and non-European ancestries using publicly available reference data (i.e., ALFA, gnomAD and ExAC); and (iii) the ancestry of participants in the studies which are listed as providing supporting evidence for the inclusion of these variants in PharmGKB.

## Methods

### Search strategy

Commercial pharmacogenomic tests were identified via Google and PubMed searches between June and August 2021 using the search terms “mental health” and “pharmacogenomics”. A pharmacogenomic test was selected if the company specifically advertised on their website that their PGx array was designed to guide selection or dosage of medication for mental health. The online search identified 14 companies that advertised PGx testing for mental health in Australia and the US. Next, companies were contacted via email and informed that we were conducting a study on ‘which alleles are most frequently tested in PGx services’, then asked to disclose the full list of the rsID identifiers of SNPs included in their array. Seven companies disclosed the manifests of the SNPs included in their arrays. We created a dataset including any genetic variant that was tested for in at least one commercial PGx array. This initial dataset contained 220 rsIDs, linked to 57 genes.

### Data analysis

Allele frequencies for each SNP were obtained, when available, from the Allele Frequency Aggregator^1^ (ALFA), the genome Aggregation Database^2^ (gnomAD), and the Exome Aggregation Consortium^2^ (ExAC) as compiled in dbSNP^16^ as of 23 November 2021. Clinical annotations for 184 SNPs were obtained from Pharmacogenomic Knowledge Base^4^ (PharmGKB). Sub-analyses related to mental health phenotypes were performed for a subset of PharmGKB entries. The full list of PharmGKB phenotypes categorized as relevant for the present study can be found in Supplementary Table 2. Data handling and analysis were performed in R^17^ using tidyr^18^ and dplyr^19^.

### CYP450 phenotype prediction and CPIC guidelines

We obtained data regarding CYP450 star alleles and HLA from the 1000 Genomes Project phase 3 release^20^ using the VCF to PED Converter online tool. Data wrangling and descriptive analysis were performed using R^17^. CPIC guidelines were available for *CYP2B6, CYP2C19* and *CYP2D6*. Gene phenotypes were estimated as follows (see Supplementary Table 7 for complete list of scores and references):

#### CYP1A2

We assigned activity values (AV) to each *CYP1A2* allele based on their functionality following the values proposed by Saiz-Rodríguez, et al. ^21^ (see Table S7). Wild alleles were assigned a value of 1. Variant rs56107638 (*7, decreased function) was missing from 1000G (GRCh37) data and for calculation purposes, all individuals were assumed to carry a *1/*1 allele. The AV for a genotype is calculated as the sum of the values assigned to each allele (e.g. *CYP1A2* *1C/*1C genotype has AV=1) and it ranged from 8.5-11. AV were translated into a comprehensive phenotype to simplify the gene association analysis as follows: ultrarapid metabolizers (AV=11); rapid metabolizers (AV=10.5); normal metabolizers (AV=10); and poor metabolizers (AV<10). There are currently no CPIC guidelines for phenotype groups in *CYP1A2*.

#### CYP2B6

Allele functionality for *CYP2B6* was based on CPIC guidelines. In the absence of a CPIC guideline for activity values or another publication where information regarding AV for CYP2B6 star alleles included in commercial PGx arrays, and with the intention to explore cross-ancestry differences using publicly available reference panels, *CYP2B6*\*4 (increased-function allele) was assigned an activity value of 1.5, *CYP2B6*\*5 (normal-function allele) was assigned an activity value of 1, and *CYP2B6*\*6 (decreased-function allele) was assigned an activity value of 0.5 (shown in Table S1). Then AV for each individual was calculated as the sum of the values assigned to each allele. Cases were categorized as ultrarapid/rapid metabolizers (AV>=6.5, or two increased-function alleles and no decreased-function alleles); normal metabolizers (AV=6; no increased-function alleles but no decreased-function alleles); intermediate metabolizers (AV=5.5, or one decreased-function allele); and poor metabolizers (AV==5, or two decreased-function alleles but no increased-function alleles). However, variant rs2279343 (*4, increased function) was missing from 1000G (GRCh37) data and for calculation purposes, all individuals were assumed to carry a *1/*1 allele.

#### CYP2C19

AV for *CYP2C19* were assigned to each simulated participant following the classification by Mrazek, et al. ^22^ (see Table S7). Wild alleles were assigned a value of 1. AV for each individual was calculated as the sum of the values assigned to each allele and they were classified as follows: ultrarapid metabolizers (AS=9, or two increased-function alleles and no no-function alleles); rapid metabolizers (AV=8.5, or one normal-function allele and one increased-function allele); normal metabolizers (AV=8, or no increased-function nor decreased-function alleles); intermediate metabolizers (AV=7 and AV=7.5, or individuals with either one normal-function allele and one no-function allele; and poor metabolizers (AV<7, or individuals with two no-function alleles. The predicted metabolizer phenotype for the combination of a no-function allele and an increased-function allele is a provisional classification. The available evidence indicates that the *CYP2C19*\*17 increased-function allele is unable to completely compensate for the *CYP2C19*\*2 no-function allele^23^.

#### CYP2C9

Individuals who carried any copies of the decreased-function alleles *CYP2C9*\*2 or *CYP2C9*\*3 were classified as poor metabolizers. Otherwise, they were classified as normal metabolizers^24^. There is no CPIC guideline for *CYP2C9* phenotypes. For the sake of completion, activity values were obtained from the CPIC website^25^ (Table S7).

#### CYP2D6

The highest functioning *CYP2D6* allele predicts in an individuals’ genotype predicts their phenotypic activity (e.g., a normal-function allele and a decreased-function results in a normal metabolizer phenotype)^26,27^.

Variant rs28371735 is missing from 1000G (GRCh37) data. Three genomic positions (42524175, 42524243 and 42525085) in 1000G data did not match dbSNP155 (GRCh37) search coordinates for query variant rs5030656, rs35742686 and rs5030655, respectively. Individuals were categorized as normal metabolizers if they had a copy of the *CYP2D6*\*2 allele. In the absence of a copy of *CYP2D6*\*2 but the presence of a decreased-function allele (i.e., *10, *17, *14, or *41), individuals were categorized as intermediate metabolizers. When only no-function alleles were present (i.e., *4 or *7), participants were categorized as poor metabolizers. This method led to an unrealistic absence of poor metabolizers in our simulated dataset. Structural variants were not considered for *CYP2D6* or any other gene. Thus, we were not able to call star alleles with whole gene deletions (*CYP2D6*\*5), or duplications (e.g., *CYP2D6*\*1×2), among others. This limits the assignment of *CYP2D6* function and phenotypes since we are not able to determine *CYP2D6* increased function alleles and therefore ultrarapid metabolizers and potentially miss no function alleles. For the sake of completion, activity values were obtained from the CPIC website^25^ (Table S7).

## Results

Our online search identified 14 commercial genetic testing services offering PGx testing specifically for mental health traits in Australia and the US (see Methods). On request, seven of these testing services provided the manifests of their arrays. In total, 220 SNPs mapping to 57 genes were included in at least one of these commercial PGx arrays (see Supplementary Table 1). Only 11.7% of the SNPs, and 6 out of 55 genes, were present on all seven arrays. All arrays included genetic variants in *CYP2A2, CYP2C19, CYP2C9, CYP2D6, CYP3A5* and *COMT*. The minimum allele set for psychiatric PGx testing proposed by Bousman et al^5^ was covered by all arrays when it came to CYP450 genes. However, only one array tested for *HLA-A* (rs1061235), one array included *CYP2D6*1* (rs28371735) and no array interrogated *HLA-B*. The lack of coverage of *HLA-A* and *HLA-B* has not changed since the 2020 review by Fan and Bousman ^13^. However, in a positive change since the review of Bousman, et al. ^12^ in 2017, there was consensus on which *CYP2D6* and *CYP2C19* star alleles were included across the arrays (Table 1 and Supplementary Table 1). From this we agree with the review by Fan and Bousman ^13^, that commercial pharmacogenomic tests are suitable, in principle, to implement most of dosing guidelines developed by the Clinical Pharmacogenetics Implementation Consortium^28^ (CPIC) relevant to psychiatry. Consensus beyond CYP450 star alleles was almost non-existent (see Supplementary Table 1).

**Table 1.** List of genetic testing services that provided information about the composition of their mental health PGx arrays, number of SNPs included (N), and genes from the minimum set for PGx in psychiatry^5^ that were not included on the arrays.

| Company | N | *CYP2C19* | *CYP2D6* | Not included<sup>5</sup> |
| --- | --- | --- | --- | --- |
| Array 1 (AUS) | 66 | *2,*3,*4,*17 | *2,*3,*4,*6,*7,*9,*10,*14,*17,*41 | *HLA-A, HLA-B* |
| Array 2 (US) | 147 | *2,*3,*4,*17 | 2,*3,*4,*6,*7,*9,*10,*14,*17,*41 | *HLA-B* |
| Array 3 (US) | 57 | *2,*3,*4,*17 | 2,*3,*4,*6,*7,*9,*10,*14,*17,*41 | *HLA-A, HLA-B* |
| Array 4 (US) | 77 | *2,*3,*4,*17 | 2,*3,*4,*6,*7,*9,*10,*14,*17,*41 | *HLA-A, HLA-B* |
| Array 5 (US) | 77 | *2,*3,*4,*17 | 2,*3,*4,*6,*7,*9,*10,*14,*17,*41 | *HLA-A, HLA-B* |
| Array 6 (AUS, US) | 66 | *2,*3,*4,*17 | 2,*3,*4,*6,*7,*9,*10,*14,*17,*41 | *HLA-A, HLA-B* |
| Array 7 (AUS, US) | 56 | *2,*3,*4,*17 | 2,*3,*4,*6,*7,*10,*14,*17,*41 | *HLA-A, HLA-B* |

Notably, 36 out of the 220 SNPs included on the arrays did not have any clinical annotations in PharmGKB. Of the remaining 184 SNPs with a clinical annotation, only 81were associated with a mental health phenotype (see Supplementary Table 2 for list of PharmGKB phenotypes considered as mental health relevant in this study). Regarding levels of evidence, only 23 out of the 81 SNPs with a mental health annotation had at least one annotation with a high level of evidence (i.e., 1A or 1B), and 80 out of 184 when considering any phenotype. Among genes associated with psychoactive medication, only *CYP2C19, CYP2C9, CYP2D6*, and *HLA-A* had clinical annotations with a high level of evidence (see Figure 1 below and Supplementary Table 1 for details on which SPNs were included in every step).

**Figure 1.**
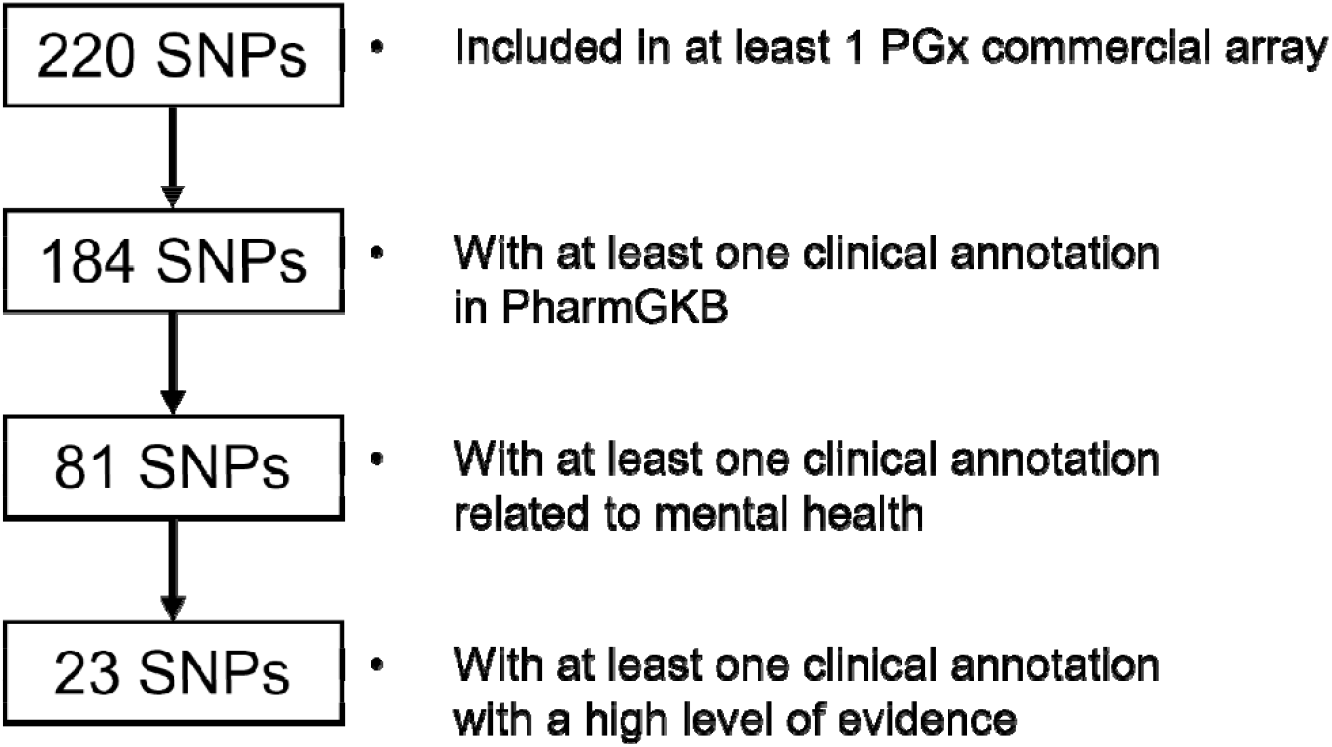
Flowchart summarizing number of SNPs originally identified, relationship with mental health phenotypes, and level of evidence.

Allele frequencies for all SNPs included in the seven commercial PGx arrays were obtained from ALFA, ExAC, and gnomAD reference panels as aggregated by dbSNP^16^ as of 11^th^ November 2021. All 81 SNPs associated with mental health phenotypes were included in ALFA. ExAC did not have frequency information for 33 SNPs, and 15 SNPs were not present in GnomAD. The largest average differences between ancestries across all pharmacogenomic variants were found between European and Asian samples (average difference in allele frequency=0.029) and African samples (0.025). The five genes with a SNP showing the largest difference in allele frequencies between European and Asian samples were *CYP3A5* (rs776746, 0.331), *ADRA2A* (rs1800544, 0.312), *GNB3* (rs5443, 0.238), *GRIK4* (rs1954787, 0.229) and *DRD3* (rs167771, 0.23). Among CYP450 genes, *CYP2C19* (N=4, 0.054) and *CYP2D6* (N=12, 0.052) showed comparatively lower average differences in allele frequency across all ancestries. The same genes showed similar average differences in gnomAD. However, 33 out of the 57 SNPs were not present in ExAC and comparison of allele frequencies across genes is not straightforward. In addition, 13 out of the 30 SNPs tested for the cytochrome P450 genes are predicted to be monomorphic by ancestry. Allele frequencies for all variants in the ALFA reference panel can be found below in Table 3 (see Supplementary Table 3-5 for allele frequencies in ExAC and gnomAD).

**Table 3.**
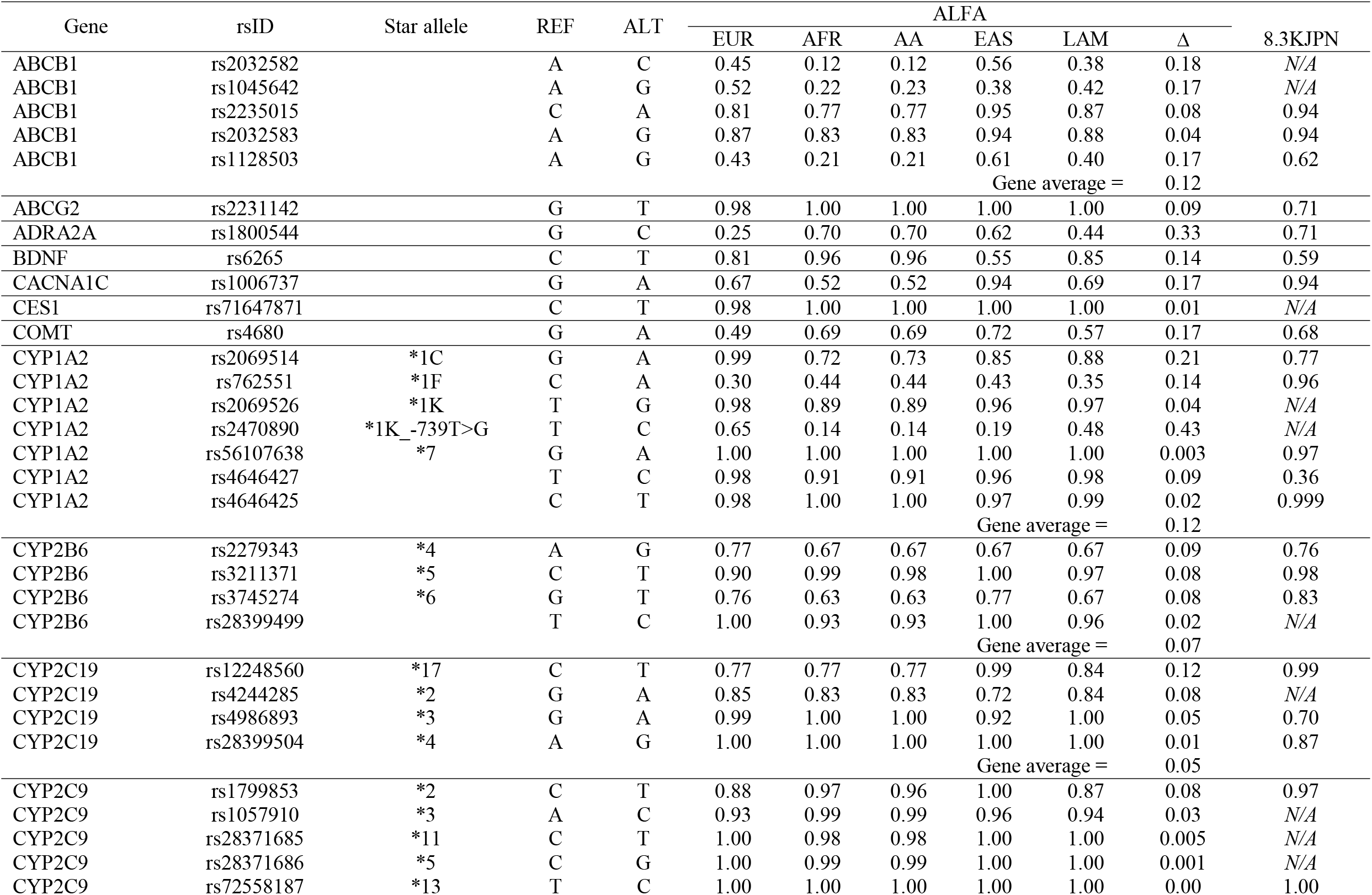
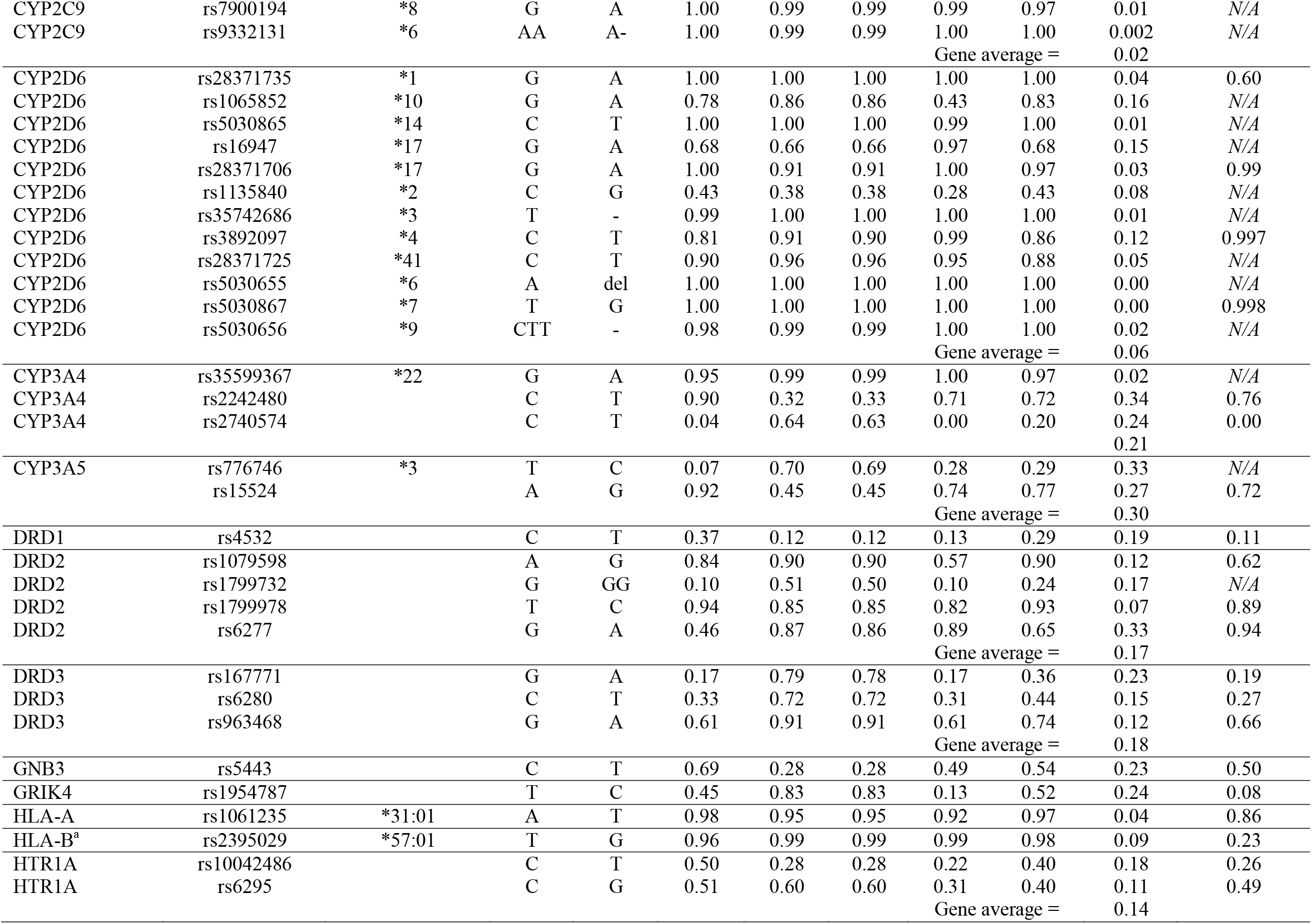
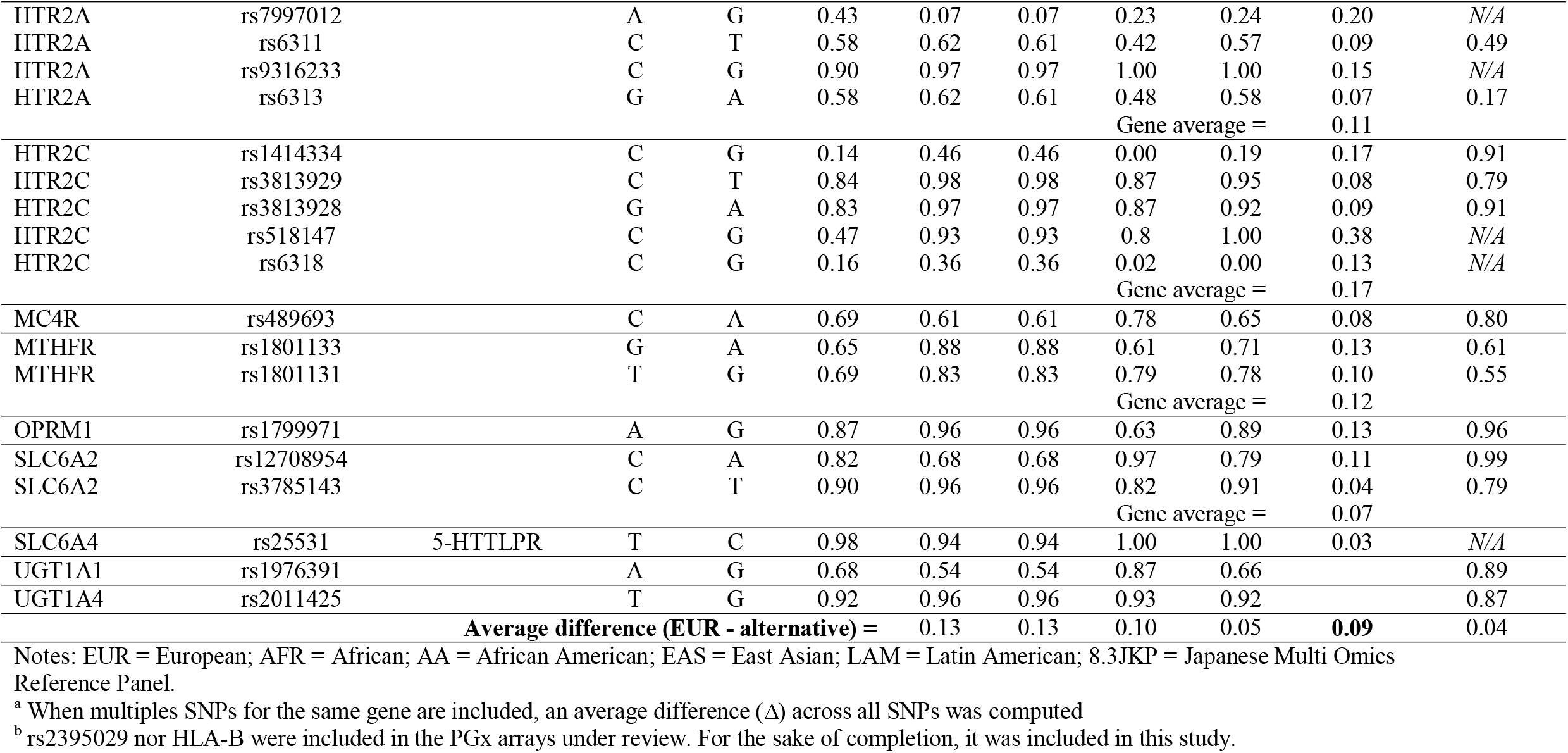
Allele frequency difference between 5 selected global populations for the 81 SNPs included in commercial PGx arrays with a clinical annotation related to mental health traits. Note: average difference (Δ) computed across all available non-European populations in ALFA (N=10).

Furthermore, *CYP2D6* or *CYP2C19* clinical decision support (CDS) guidelines are available for sertraline, escitalopram, duloxetine, fluoxetine and venlafaxine, which are among the top 50 most prescribed medications annually in the US^29^ and Australia^30^. We see that *CYP2C9*2* (rs1799853), associated with normal CYP2C9 function, is more common in European and Latin American populations but less common in African East Asian populations. *CYP2D6*4* (rs3892097), a loss-of-function allele that results in a nonfunctional CYP2D6 protein^25^, is also more common in European populations but far less common in African populations and very rare in Asian populations. In another example, *CYP2C19*17* (rs12248560) results in enhanced transcription and an ultra-rapid metabolizer phenotype and is an actionable genotype^28^. *CYP2C19*17* is relatively common in European and African populations but very rare in Asian populations. Consequently, differences in allele frequency influence the applicability of CDS for individuals with non-European ancestry given that the proportion of individuals categorized as carrying a high-risk gene phenotype would change and therefore the benefit of PGx testing for different world populations. With the aim of estimating the utility of psychiatric PGx testing using commercial arrays across ancestry, first we downloaded genotype data for SNPs in the CYP450 genes included in commercial arrays from 1000 Genomes Project^20^. Next, we calculated the proportion of individuals that would be considered to have a high-risk genotype according to CPIC guidelines for the African, Ad Mixed American, East Asian, European and South Asian macro-populations. Summary of results can be found below in Table 4.

**Table 4.**
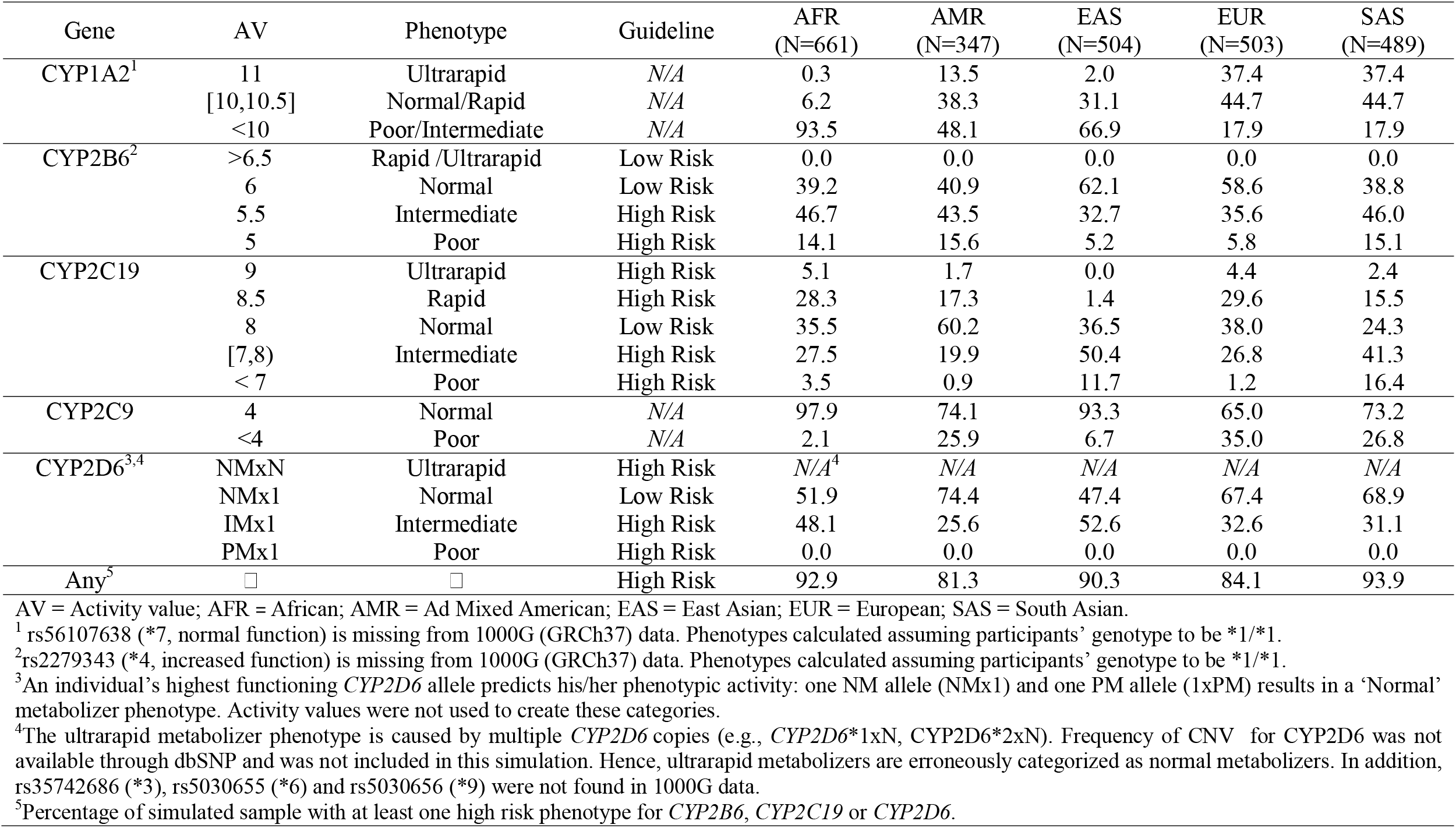
Predicted phenotype frequencies for cytochrome P450 genes using 1000 Genomes Project data for five macro-populations.

Based on the haplotype frequencies observed in the 1000 Genomes cohort, we could expect PGx testing to be as useful in identifying non-European individuals that could benefit from genetically-guided dosing and medication selection as they are for European samples (see Figure 3). While informative, our results cannot be used in any case for public recommendations and further evaluations of haplotype frequencies in larger non-European samples should be conducted. Additional information regarding activity values and gene phenotype prediction can be found in Online Methods.

Finally, in 2019, PharmGKB recommended the standardization of ancestry groups reported in PGx studies using seven geographically defined groups (i.e., American, Central/South Asian, East Asian, European, Near Eastern, Oceanian, and Sub-Saharan African), plus two admixed groups (African American/Afro-Caribbean and Latino)^31^. Currently, each clinical annotation aggregated by PharmGKB includes metadata about the biogeographical origin of the sample from the original study. We extracted this information and analyzed biogeographical origin of supporting evidence for clinical annotations associated with mental health phenotypes, organized by level of evidence. Our review showed that across all genes with any clinical annotation, regardless of their level of evidence, 25 out of 46 annotations originated from European samples, 11 from samples with unknown ethnicity, 9 from East Asian samples, and 1 from an ad-mixed sample. Out of the 8 clinical annotations with a high level of evidence for *CYP2C19* in mental health phenotypes, 4 were based on European samples, 3 from samples with unknown ethnicity, and 1 from an East Asian sample. In the case of *CYP2D6*, out of the 25 well-supported clinical annotations, 16 were based on European samples, 5 from samples with unknown ethnicity, and 4 from an East Asian sample. For both *CYP2C19* and *CYP2D6*, no annotations with a high level of evidence were determined in studies of African samples. The breakdown of biogeographical origin of supporting evidence in PharmaGKB for clinical annotations among the five cytochrome P450 genes most relevant for psychiatry can be found in Figure 2 and Supplementary Table 6.

**Figure 2.**
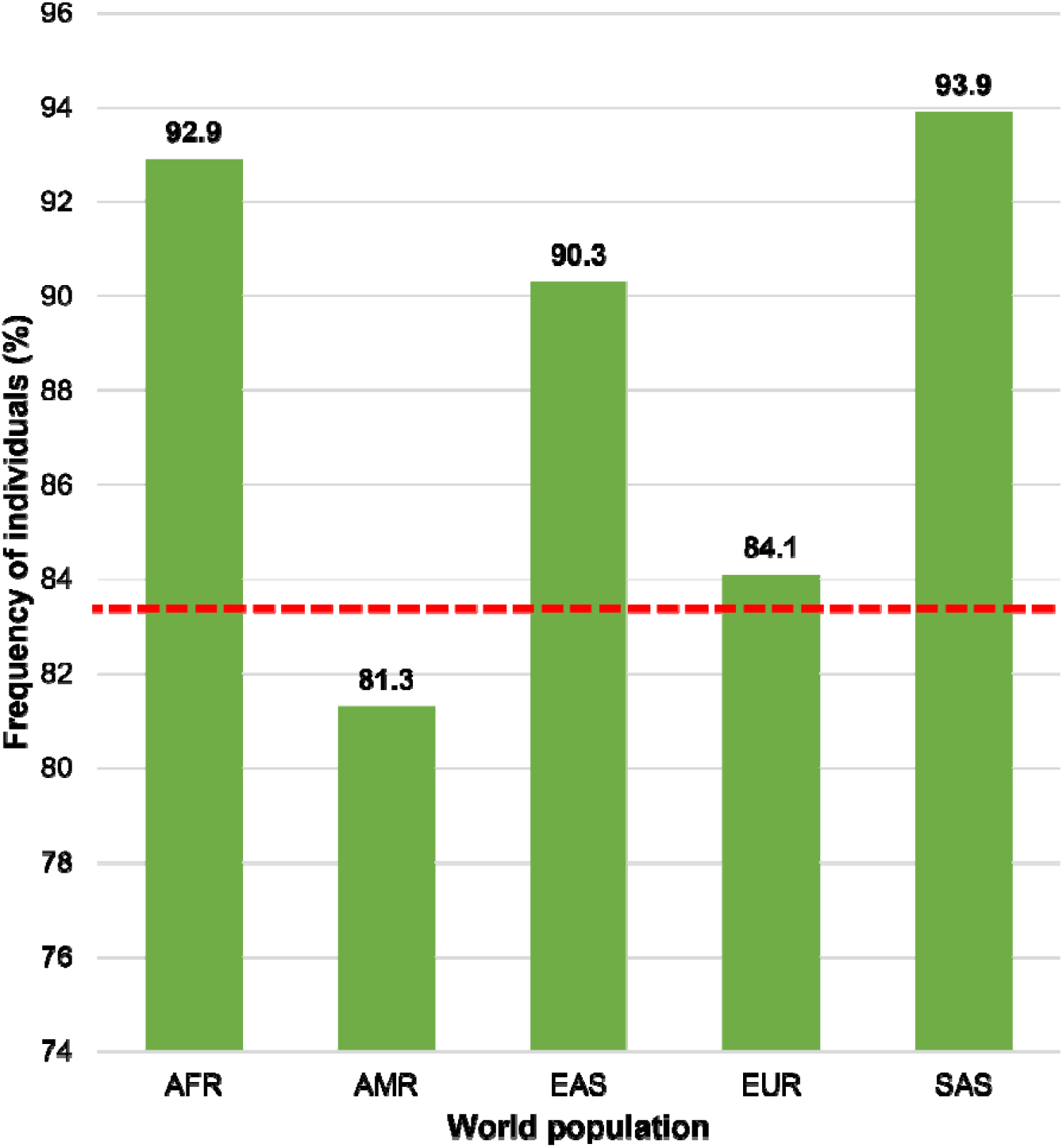
Expected frequency of individuals with at least one high risk genotype according to CPIC guidelines based on 1000 Genomes Project data. Note: AFR = African; AMR = Ad Mixed American; EAS = East Asian; EUR = European; SAS = South Asian.

**Figure 3.**
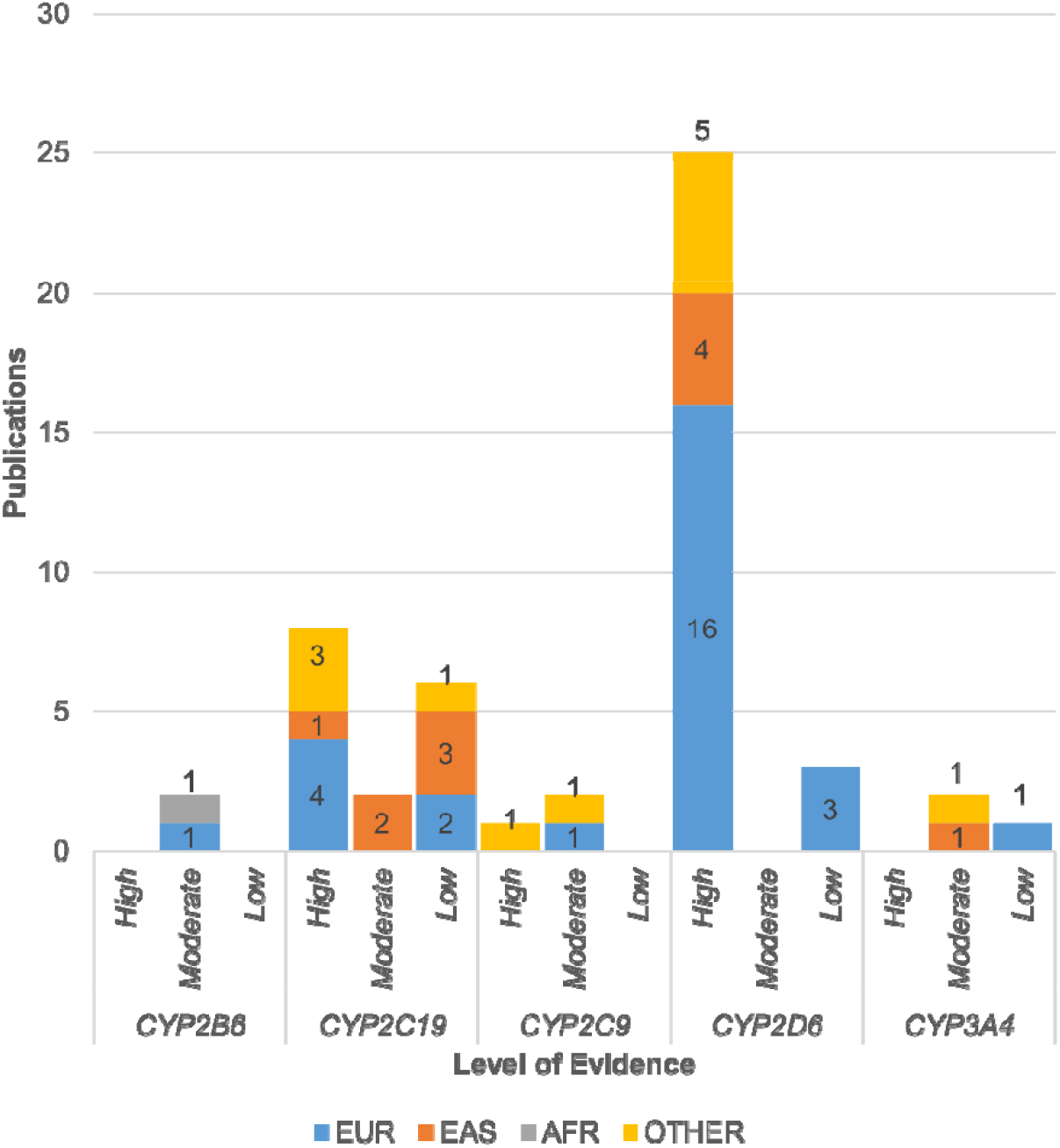
Number of supporting studies for clinical annotations in PharmGKB organized by cytochrome P450 gene and level of evidence. In summary, out of 46 gene-drug supporting studies associated with mental health phenotypes, 25 only included European samples, 11 included samples of unknown ethnicity, 9 included East Asian samples, and 1 ad-mixed sample.

In summary, the frequencies of genetic variants included on these PGx arrays differs significantly between European and non-European populations based on public reference data, and most of the supporting evidence for clinical annotations in PharmGKB originated from European or unknown samples. The over representation of European populations in genetic research is well known^9^ and remains one of the key barriers to the translation of clinical recommendations in pharmacogenomics in genetically diverse communities. Unless addressed, this will result in an increased health disparity in precision medicine. In addition, critical population-specific variants might also be overlooked if translation efforts are continued without including non-European samples, and future studies might identify new drug targets in populations where key pharmacogenomic variants are more prevalent, among other reasons^6^.

The present study did not evaluate how the seven commercial PGx tests were advertised, or if they included disclaimers regarding the applicability of their results to individuals of different ancestries. In our searches for online commercial PGx testing product information we did not find information regarding differences in allele frequency across ancestries. While this omission might be a consequence of systematic bias towards European population, it is also the case that the present study suggests that consumers from non-European backgrounds would benefit the same if not more from this technology. Nevertheless, the validity of CDS based on PGx arrays across diverse populations also needs to be examined before PGx testing can be considered standard of care. While frequency of high-risk phenotypes is high among all world populations, impact of PGx-guided treatment still will requires large studies including economic, health service use and psychological data to prove its utility and avoid hype.

Furthermore, our findings regarding the number of SNPs that were included in the PGx arrays for mental health that either had no clinical annotations for mental health phenotypes or no clinical annotations in publicly available data sources, raises the question of what evidence was used to support the inclusion of these variants on the array and the extent to which these variants are used in PGx reports provided to clinicians and health professionals.

There are several limitations to be noted. First, our online search strategy may have missed PGx testing companies that did not appear in our searches. Second, we were only able to access manifests for 7 commercial PGx arrays and therefore generalization from this study is limited. Third, we limited our description of high risk genotypes to 1000 Genomes cohort which is relatively small and was not designed to evaluate pharmacogenomic profiles. Future studies should include genotype data from large world population cohorts and evaluate heterogeneity in “measured” phenotypes in order to accurately estimate the utility of current PGx arrays in mental health for non-European populations. In conclusion, the present study synthesizes key barriers to translation of PGx services in diverse populations and highlights current needs in supporting evidence for gene-drug interactions for contemporary personalized psychiatry.

## Data Availability

All data produced in the present study are available upon reasonable request to the authors.

## Acknowledgements

This study was funded by the Medical Research Future Fund (APP1200644). J.J.M. was supported by MRFF APP1200644. S.E.M. is supported by a National Health and Medical Research Council Investigator Grant (APP1172917). We would like to thank Stanford University and the PharmGKB resource or providing the computational resources that contributed to this study.

## Author contributions

J.J.M., K.S. P.A.L., G.P., and S.E.M. contributed to all aspects of the paper. J.J.M. wrote the manuscript and analyzed the data. K.S. retrieved the data from dbSNP. P.A.L., G.P. and S.E.M. provided critical revisions to the manuscript. P.A.L., G.P. and S.E.M. secured the funding for the project.

## Additional information

The authors declare no competing financial interests. Correspondence and requests for materials should be addressed to J.J.M.

